# Cumulative COVID-19 incidence indicators should account for the population-at-risk dynamics

**DOI:** 10.1101/2022.10.18.22281225

**Authors:** Pere Masjuan, Sergi Trias-Llimós, Camilo Rojas, Arul Prakash, Pedro Gullón, Aurelio Tobias, Antonio López-Gay

## Abstract

Risk estimators for COVID-19 propagation based on the incidence rate of new cases can be misleading as they usually fail to account for the fraction of population immunized by infection or vaccination.This misconception yields different incidence rates, as we illustrate using the daily number of COVID-19 reported cases in Spain during the pre-vaccine period, between 15/01/2020 and 11/07/2021. An increase in the incidence rate of about 7% is found when properly accounting for the population at risk. Our results demonstrate that accounting for dynamic changes to the immunized fraction of the population is necessary for accurate risk estimation. We hope that our findings can lead to more effective strategies for pandemic response.

---

Risk estimators for the COVID propagation are essential for assessing the real risk to the population posed by the different COVID waves (1). These estimates usually rely on the 14-day cumulative incidence rate of new COVID-19 cases (IR_14_ in the following), defined as the cumulative number of reported COVID-19 cases over 14 days divided by the population at risk per 100,000, following the classical epidemiological definition of incidence rate (2):

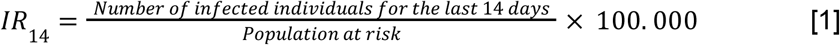

The IR_14_ metric has been particularly useful to monitor the dynamics of the pandemic as well as to set thresholds used to trigger mitigation strategies during the pandemic (3). A common assumption within COVID-19 risk estimates has been to use the *total* population as the population at risk in Eq.[1]. However, the risk of being infected should take into consideration those who were not yet infected instead of the total population as the population at risk. Failing to account for the dynamics of the exposure population could, importantly, bias downwards real transmission levels. This bias may delay mitigation policies and complicate the understanding of the pandemic’s evolution, as it prevents a correct interpretation of the comparison of IR_14_ over time (comparing levels of different waves) and across space (comparing the same waves across countries or regions regardless of the unique previous evolution of the pandemic in each area). This correction is especially important during the first waves of the COVID-19 pandemic when reinfection was nearly unattainable (4).

We propose to correct the IR_14_ by accommodating the estimates to the situation where a non-negligible part of the population was not exposed to the risk (already immunized in the first pandemic stages). In doing so, we take into account that the *exposed-to-risk* population (those who can get infected) is lower than the total population when part of it has already had the infection. Holding constant the numerator, a reduction of the denominator represents an overall higher risk.

To illustrate this potential bias, we collected the daily number of COVID-19 reported cases in Spain during the pre-vaccine period, between 15/01/2020 and 11/07/2021, and calculated the IR_14_ for each day using the total Spanish population as a denominator. Within the pre-vaccine period (between 15/01/2020 and 11/07/2021), we assume the infection provided 6, 8, or 10 months of immunity after the disease is contracted, which is a conservative assumption in the first stages of the pandemic (4). We do so by modifying the IR_14_ as:

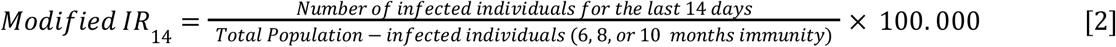

We observed that the population at risk as defined in our formula was decreasing over time as COVID-19 infections were increasing, and represented around 93% of the total Spanish population in Spring 2021 (Figure 1) when accounting for a 10-month immunity period.

**Figure 1.**
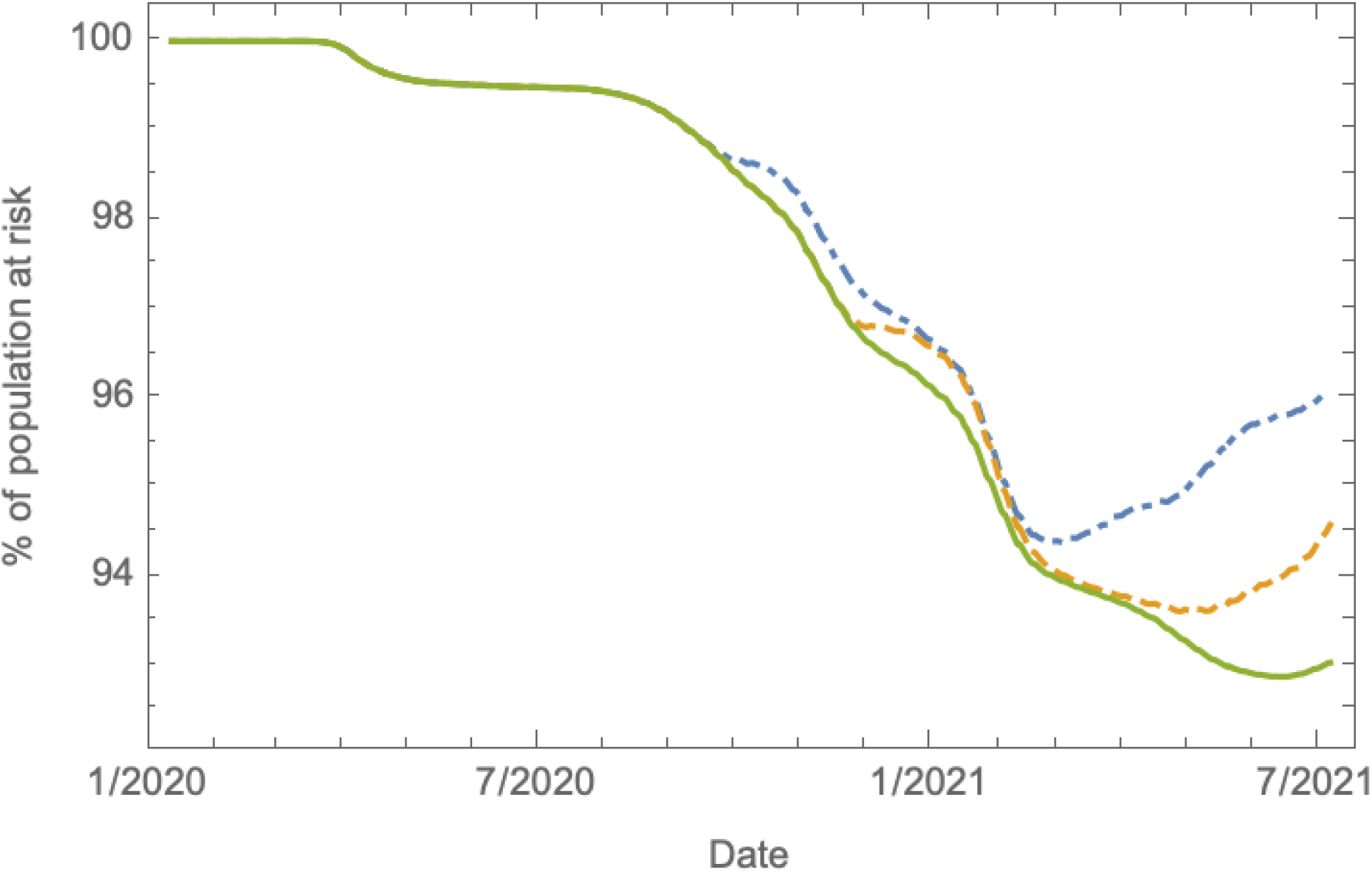
Percentage (%) of the population at risk of COVID-19 infection relative to the total Spanish population during the pre-vaccine period, from 15th January 2020 to 11th July 2021, assuming the immunity after the disease is contracted may last for 6 (dot-dashed), 8 (dashed), 10 (solid) months.

**Figure 2:**
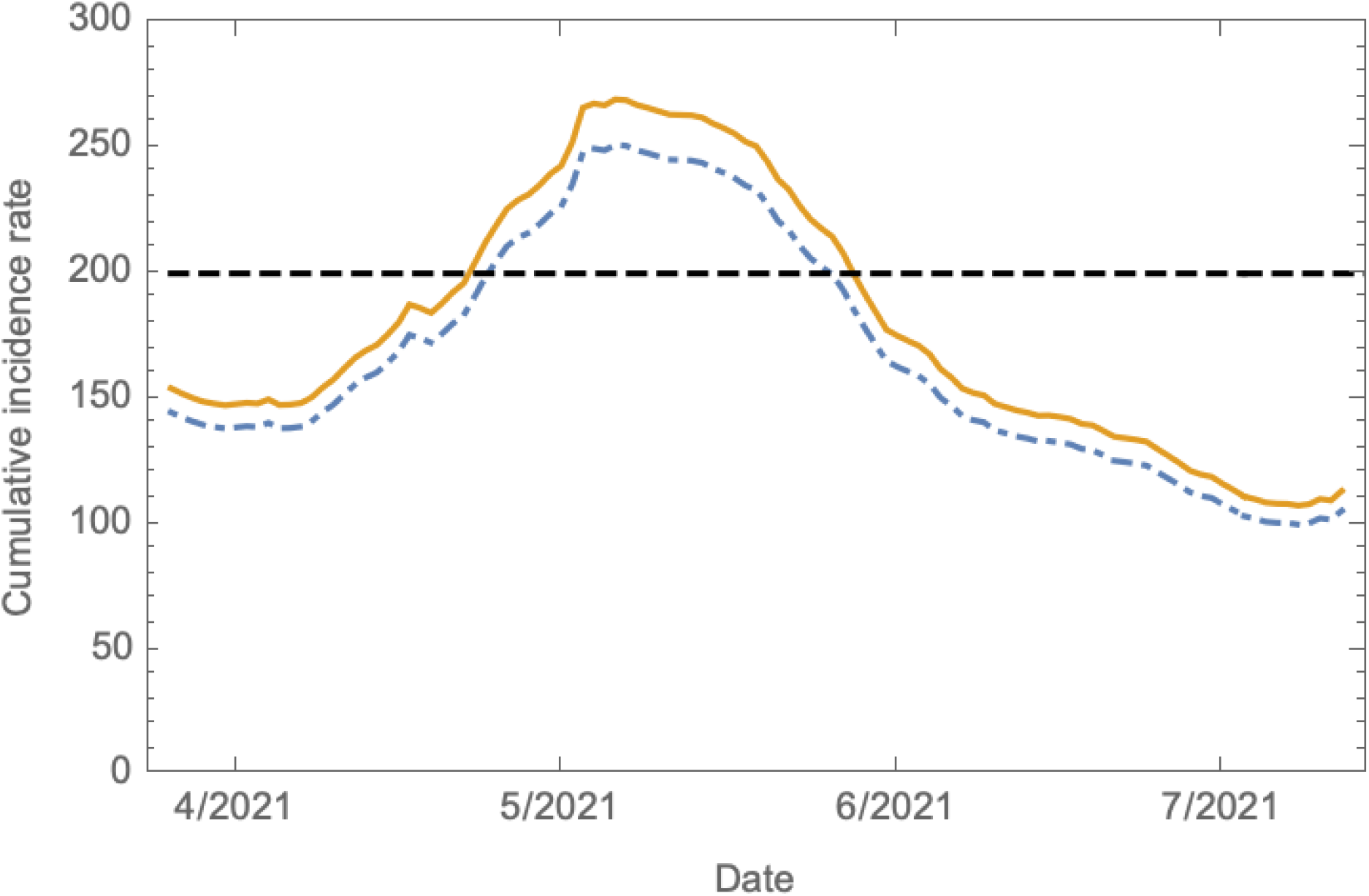
14-day cumulative incidence rate of new COVID-19 cases calculated using Eq.[1], dot-dashed line, compared to its modified version calculated using Eq.[2], solid line, assuming 10 months of immunity. The threshold of 200 new cases is shown with a dashed line.

This decline in the denominator represents, as expected, a higher incidence rate of new COVID-19 cases, particularly in Spring 2021 for the Spanish case. For example, our estimates suggest that assuming 10 months of immunity, the threshold of 200 new COVID-19 cases per 100,000 inhabitants surpassed on 10/4/2021 would have been surpassed 5 days earlier if the population at risk only accounted for those individuals at risk of infection.

Our approach could be considered conservative as not all cases were reported, especially during the first wave, when there was a significant lack of testing for asymptomatic and mildly symptomatic individuals (5). We estimated that around 3-4% of the population had been infected by December 2020, which contrasts with higher estimates based on a seroprevalence study (9.9%) (6). Therefore, within the pre-vaccine period, using the total population as a denominator for the COVID-19 cumulative incidence rate represents an underestimation of the actual COVID-19 transmission risk.

We only considered the pre-vaccine period since we acknowledge vaccines could have altered significantly the population at risk, decreasing it with the massive vaccination plans and then increasing it with the subsequent loss of immunity after several months (7). Furthermore, the risk of reinfection also increased substantially over time with the appearance of new variants (8,9). These factors can potentially significantly impact both the population at risk and the rate of new COVID-19 cases and will need to be carefully modeled in further studies.

In conclusion, correcting the incidence rate of new COVID-19 cases by the fact that the population at risk is not the total population, but a changing fraction of it provides higher and more accurate estimates of COVID-19 risk. Therefore, the population at risk should be carefully considered when monitoring epidemic data.

The data underlying this article come from the Centro Nacional de Epidemiología and the Instituto de Salud Carlos III, and are available at https://rubenfcasal.github.io/COVID-19/.

## Data Availability

The data underlying this article come from the Centro Nacional de Epidemiologia and the Instituto de Salud Carlos III, and are available at

https://rubenfcasal.github.io/COVID-19/

## Acknowledgment

The authors affiliated to CED acknowledge the support of the CERCA institution, Centres de Recerca de Catalunya. AL acknowledges the support from the Talent Research Program (Universitat Autònoma de Barcelona). PM, CR, and AP acknowledge the support from the Ajuntament de Barcelona. STL acknowledges research funding from the Juan de la Cierva-Formación (Spanish Ministry of Science and Innovation, FJC-2019-039314-I). PG is supported by Instituto de Salud Carlos III (ISCIII) and co-funded by the European Union (PI18/00782 and PI21/01868) and by Comunidad de Madrid (Línea de actuación “Estímulo a la Investigación de Jóvenes Doctores”, CM/JIN/2021-028). AT was supported by MCIN/AEI/10.13039/501100011033 (grant CEX2018-000794-S).

